# Disparities in Autonomy during Pregnancy in Canada: Findings from the national RESPCCT Study

**DOI:** 10.1101/2025.08.01.25332643

**Authors:** Kathrin Stoll, Karen Hodge, Bhavya Reddy, Raymonde Gagnon, Régine Tremblay, Sylvie Lévesque, Wanda Phillips-Beck, Nisha Malhotra, Rohan D’Souza, Ali Tatum, Saraswathi Vedam

## Abstract

**Background:** Very little is known about how childbearing people in Canada experience decision-making during pregnancy, and whether experiences of autonomy differ by personal characteristics and context of care.

**Methods:** Using national survey data, we examined associations between autonomy (measured with the validated My Autonomy in Decision-Making (MADM) scale and 1) participant characteristics, and 2) contextual factors including timing of prenatal care initiation, and length of appointments. We calculated the odds of high autonomy and 95th confidence intervals among subgroups, controlling for pregnancy year, repeat observations, and gravidity. Analyses were stratified by provider type (midwives or physicians).

**Findings:** A total of 5389 participants reported 7049 interactions with healthcare providers in Canada between 2009 and 2022. The odds of high autonomy were significantly lower among Indigenous (AOR=0.73; 95 % CI: 0.59-0.90) and Middle Eastern participants (AOR=0.57; 95 % CI: 0.39-0.83), among younger participants (≤25 years; AOR=0.47, 95 % CI: 0.36-0.62), and people with a disability (AOR=0.77, 95 % CI: 0.61-0.98), lower educational attainment (AOR=0.37, 95 % CI: 0.27-0.50), and insufficient income (AOR=0.47, 95 % CI: 0.36-0.62). In addition, participants who identified as recent immigrants (AOR=0.60, 95 % CI: 0.37-0.97), and people needing two or more social services during pregnancy (AOR=0.33, 95 % CI: 0.22-0.50), with high risk pregnancies (AOR=0.46, 95 % CI: 0.39-0.0,54), pre-pregnancy BMI > 30 (AOR=0.78, 95 % CI: 0.66-0.91), history of substance use disorder (AOR=0.68, 95 % CI: 0.49-0.95) and higher levels of discrimination (AOR=0.69, 95 5 CI: 0.57-0.83) were less likely to report high autonomy. Factors associated with higher odds of autonomy included midwifery (versus physician care), early entry into prenatal care, sufficient appointment time, and access to a preferred prenatal provider.

**Conclusions:** Minoritized and socioeconomic disadvantage participants reported significantly less autonomy. These differences persisted across models of care, indicating structural inequities in person-centered care during pregnancy.

## Background

The ability of childbearing people to lead decisions about their care is a reproductive right (1–3). Healthcare providers facilitate autonomy in decision-making by communicating the best available evidence about options for care using lay language, checking with patients to ensure they have understood all the information, and considering their cultural and personal preferences (4). Supporting autonomy in decision-making during perinatal care is central to providing high-quality, person-centered care (5). Violations of patient autonomy include disregarding the wishes and preferences of pregnant people, performing tests and procedures without consent, using coercion and threats, biased communication and disregarding or minimizing the knowledge of women about their own bodies or cultural practices (4,6,7). Disregard for autonomy during perinatal care is a uniquely gendered problem, where power is enacted on the bodies of women both subtly and with force (8,9). For these reasons, autonomy in perinatal care is also a core component of eliminating gender-based violence and promoting respectful care (10).

Qualitative studies illustrate that feeling a loss of control during birth can negatively affect people’s birth experience (11,12), lead to lasting trauma (12), and erode trust between patients, their providers and ultimately the health system (12–15). Emerging quantitative studies on the subject demonstrate that loss of patient autonomy is associated with postpartum mental health problems, such as depression and post-traumatic stress disorder (PTSD) (16–18). Loss of autonomy also affects health professional trainees, e.g. PTSD symptoms have been reported by midwifery students who observed loss of autonomy of women during clinical placements (19).

### Disparities in perinatal autonomy in Canada

Studies suggest there are inequities in the lived experience of autonomy, especially for pregnant and childbearing people with marginalized identities and life circumstances. Indigenous, disabled, racialized and incarcerated women have suffered disproportionately from reproductive rights violations in Canadian history and continue to report egregious violations such as forced sterilization(20). Childbearing people in Canada are treated differently based on their characteristics because of how colonialism, racism, ableism, weight stigma, and other systems of oppression privilege some and disadvantage others (1). These systems of oppressions operate simultaneously and disproportionally affect those with intersecting identities and life circumstances (21,22). In one provincial study in British Columbia (n=2051) childbearing people with medical or social risk factors, less education, and those who reported being treated with less respect by healthcare providers based on their race or ethnicity were also less likely to experience autonomy in decision-making (23). For decades, newcomers to Canada have reported that their preferences are not respected when they receive perinatal care (24). Muslim immigrant women in Newfoundland have noted that their cultural and religious needs were disrespected and when they advocated for their needs, they were met with frustration and anger from healthcare providers (25).

We hypothesized that experiences of autonomy during pregnancy are embedded in structural inequities, and affected by social position, adverse life circumstances, pregnancy factors and context of care (26,27). Previous research suggests groups who have been marginalized on the basis of Indigeneity, immigrant status, disability, gender, substance use, socio-economic status, among other social factors, may face power inequities around decision-making, as has been documented with related aspects of perinatal care experiences in Canada (28–30). Hence, using a large, national dataset of patient-reported outcomes and experiences, we designed a study to assess the drivers of disparities in autonomy during perinatal care. We also examined whether context of care, such as access to early prenatal care, enough time during prenatal appointments, and access to one’s preferred prenatal provider might reduce or eliminate inequities in autonomy during pregnancy among childbearing people.

## METHODS

### Theoretical frameworks

The data presented in this paper was collected during *The Research Examining Stories of Pregnancy & Childbirth in Canada Today (RESPCCT)* study, a multi-phase, national, community participatory action research (CPAR) project (31). Several tenets of CPAR make it a promising path to addressing inequities in experiences of perinatal care. These include a framework informed by the principles of reproductive justice and health system accountability (31–33), the centering of lived experiences of oppressed groups in every phase of research to account for power imbalances and the focus on actionable findings to reduce inequities.

The authors of the current paper position themselves as clinicians, parents, community health workers, and researchers with lived experiences as well as extensive exposure to perinatal health systems and diverse populations of women in Canada. One author is an Anishinaabe scholar with experience working and living in an Indigenous community; and other authors self-identify as minoritized by racial, immigrant status, sexual identity, and/or living with disabilities. These perspectives are embedded in our approach to the analysis and interpretation phases of the RESPCCT study (31).

### Study design and recruitment

The RESPCCT study team included a Community Steering Council (CSC), multidisciplinary investigators, community health workers, and leaders from professional associations. Using a community participatory action research approach, we conducted a systematic review and multi-stakeholder Delphi process to co-develop a survey that was translated into 7 languages and pilot tested by a representative sample of people with previous pregnancy experiences and diverse identities and circumstances. Recruitment was achieved via social media platforms, distribution by non-governmental organization (NGO) partners and professional bodies, with the help of 18 Regional Recruitment Coordinators who engaged communities that are under-represented in Canadian perinatal research. Detailed methods for item generation, survey construction, and data collection are reported elsewhere (31,34).

RESPCCT participants articulated their experiences of respectful perinatal care through 388 closed and open-ended survey items, across 17 domains of respect and disrespect during pregnancy, birth, and the postpartum period, including validated patient-reported experience measures that assess autonomy, respect and mistreatment (34–37). Socio-demographic questions ensured that respondents could self-identify their ethno-racial group, sexual and gender identity, disability status during pregnancy, socio-economic status, and other health or social factors, and life events. Other items captured provider type, mode of birth, obstetric interventions, and details on the birth and postpartum experience. The survey took approximately 60 minutes to complete and was available in eight languages (Arabic, Chinese -simplified, Chinese -traditional, English, French, Inuktitut, Punjabi, and Spanish). A screen reader version of the survey was also available. Three distinct survey pathways collected information on current and past pregnancy experiences, as well as from those reporting on pregnancy loss. People from all provinces and territories (n=6096) participated.

### Study Measures

#### Outcome Measure

Autonomy was measured with the validated 7-item *My Autonomy in Decision Making (MADM)* scale that assesses the level of agency that participants experience during discussions with their care provider about their options for care during pregnancy (35). Each MADM item has six response options (ie. strongly disagree to strongly agree) with total scores ranging from 7-42. Higher scores indicate a higher degree of autonomy. The items were introduced as follows: *Please answer the following questions to describe your discussions with your doctor or midwife DURING PREGNANCY about your options for care (for example: prenatal testing, starting your labour, medications, caesarean, place of birth, newborn care, etc.)*.

Prior to completing the scale, participants indicated the type of healthcare provider that their answers referred to: family physician (FP), midwife (MW), obstetrician (OB), nurse practitioner (NP) or ‘not applicable; I did not have a doctor or midwife’. Each participant could report on their decision-making interactions with up to two different healthcare providers in the same pregnancy by completing the scale items more than once. See Table 1 for the MADM items, stratified by the provider type that was rated. To date the tool has been translated into 20+ languages and consistently shows high internal consistency reliability across countries (18,38,39).

**Table 1:** My Autonomy in Decision-Making (MADM) Scale items and proportion of participants who agreed or strongly agreed with each item, stratified by the provider type.

|  |  | Family Doctor<br>(n=1245) | OB<br>(n=2218) | Midwife<br>(n=1938) |
| --- | --- | --- | --- | --- |
| 1 | My provider asked me how involved in decision making I wanted to be. | 485 (39.1) | 734 (33.2) | 1372 (71.0) |
| 2 | My provider told me that there are different options for my maternity care. | 442 (35.8) | 591 (26.8) | 1631 (84.7) |
| 3 | My provider explained the advantages/disadvantages of the maternity care options. | 387 (31.3) | 605 (27.5) | 1511 (78.3) |
| 4 | My provider helped me understand all the information. | 565 (45.8) | 945 (42.9) | 1608 (83.6) |
| 5 | I was given enough time to thoroughly consider the different care options. | 528 (42.8) | 797 (36.1) | 1611 (83.6) |
| 6 | I was able to choose what I considered to be the best care options. | 619 (50.4) | 957 (43.5) | 1640 (85.1) |
| 7 | My provider respected my choices. | 803 (65.2) | 1256 (57.2) | 1670 (86.6) |

*Response options: 1- Strongly disagree, 2- Disagree, 3- Somewhat disagree, 4- Somewhat agree, 5- Agree, 6- Agree Strongly*

#### Independent variables

Independent variables included personal characteristics and circumstances (ethnoracial identity, gender identity, age, education, disability and relationship status, immigration history, income, substance use during pregnancy, pregnancy risk, and pre-pregnancy BMI). We note that the ‘Indigenous’ variable for this analysis combines the three Indigenous groups in Canada—First Nations, Inuit, and Métis. We also included number of services that respondents needed during pregnancy as an indicator of social vulnerability. Options included: Welfare or social assistance, drug or alcohol treatment, mental health treatment, help to quit smoking, safe house or shelter, homeless shelter, resettlement or refugee settlement centre, family protective services, food banks, disability resources/aids, transportation assistance to access services, and legal aid. Participants could write in up to 3 additional types of services and these were also counted. We then recoded the sum into three categories: None, 1,2 or more.

We included the 9-item Day-to-Day Discrimination index (40), a tool that was intentionally designed without attribution to specific social identities because of evidence that the harmful effects of discrimination are not tied to specific reasons for discrimination (41) (Sample item: *Because of who you are, have you been called names or heard/saw your identity used as an insult?*). We also included the tool because Adkins and others warn about the use of race and other identifiers as proxies for discrimination or racism (42,43). Discrimination scores ranged from 0-18. Cut off scores for this measure have not been published, hence we recoded the continuous discrimination scores into quartiles: no discrimination (score of 0-bottom two quartiles), low discrimination (scores of 1-3) and moderate/high discrimination (scores of 4 or higher-top quartile).

To stay true to our community-driven mandate to explore modifiable factors that might reduce disparities in perinatal experiences, we also examined 1) early access to prenatal care, 2) sufficient time during prenatal appointments, and 3) access to a person’s preferred healthcare provider. In addition to exploring the association between these three variables and autonomy, we also summed the three access conditions to determine how the number of optimal access conditions related to autonomy experiences. This approach goes beyond looking at individual factors in isolation and instead considers how their combined effects create unique experiences and opportunities for action. See Additional File 1 for a description of variables that were included in the analysis.

### Stratifying by provider type

The distribution of MADM scores was different depending on whether participants reported on interactions with physicians or midwives. Respondents who were cared for by physicians reported low autonomy scores more often while midwifery clients reported high scores more often (see Figures 1 & 2). Given these different distributions, and existing literature that demonstrated differences in MADM scores by provider type (23,44–47), we opted to present bi and multivariable findings separately for scores related to interactions with physicians versus midwives. We combined family physicians with obstetricians under the category ‘physician’ since both types of physicians had similar MADM scores (see Table 1). Few people submitted MADM scores in reference to nurse practitioners; and hence, these cases were excluded.

**Figure 1:**
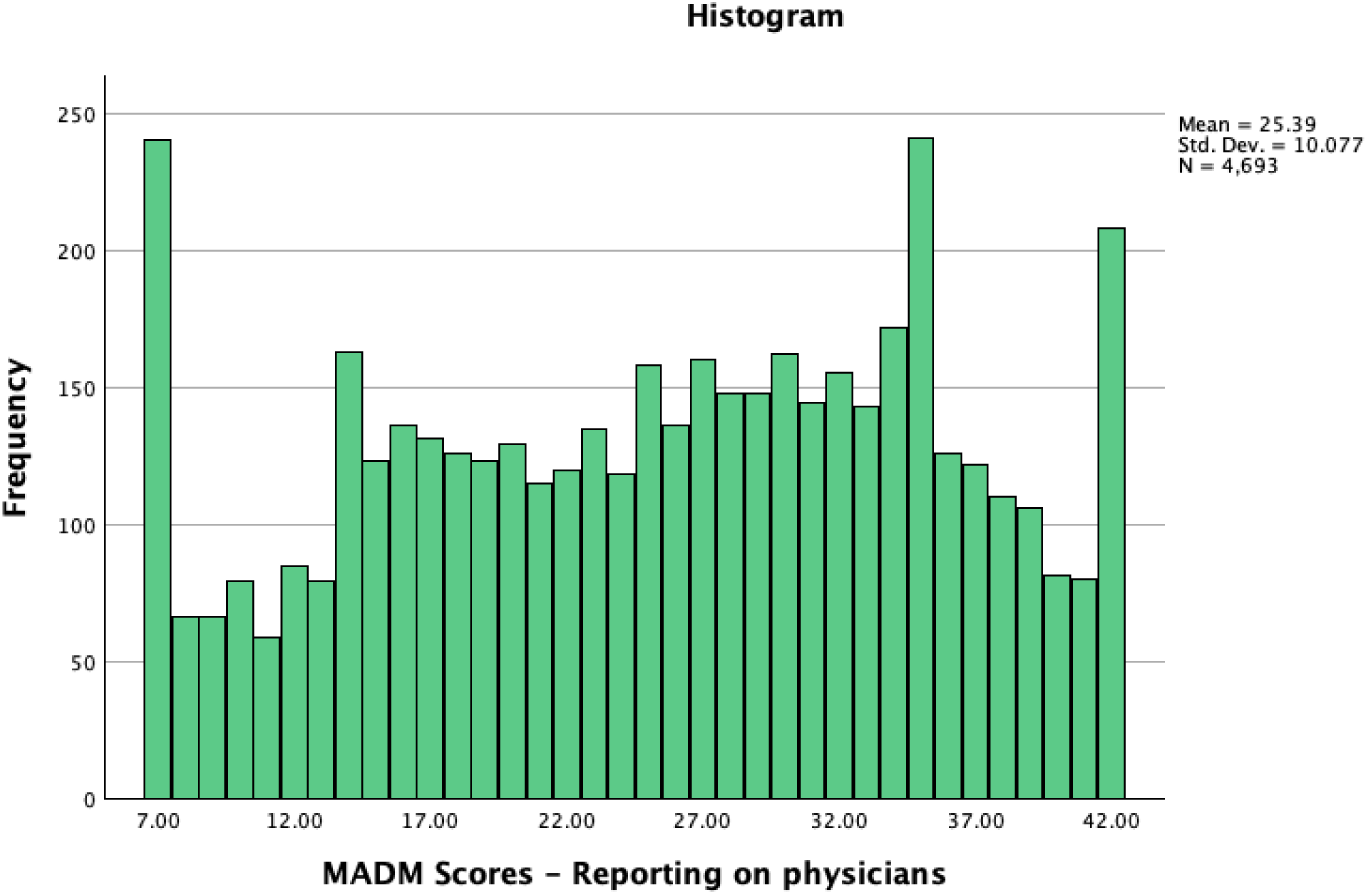
Distribution of MADM scores among participants reporting on physicians (n=3386 participants reporting 4693 autonomy experiences)

**Figure 2:**
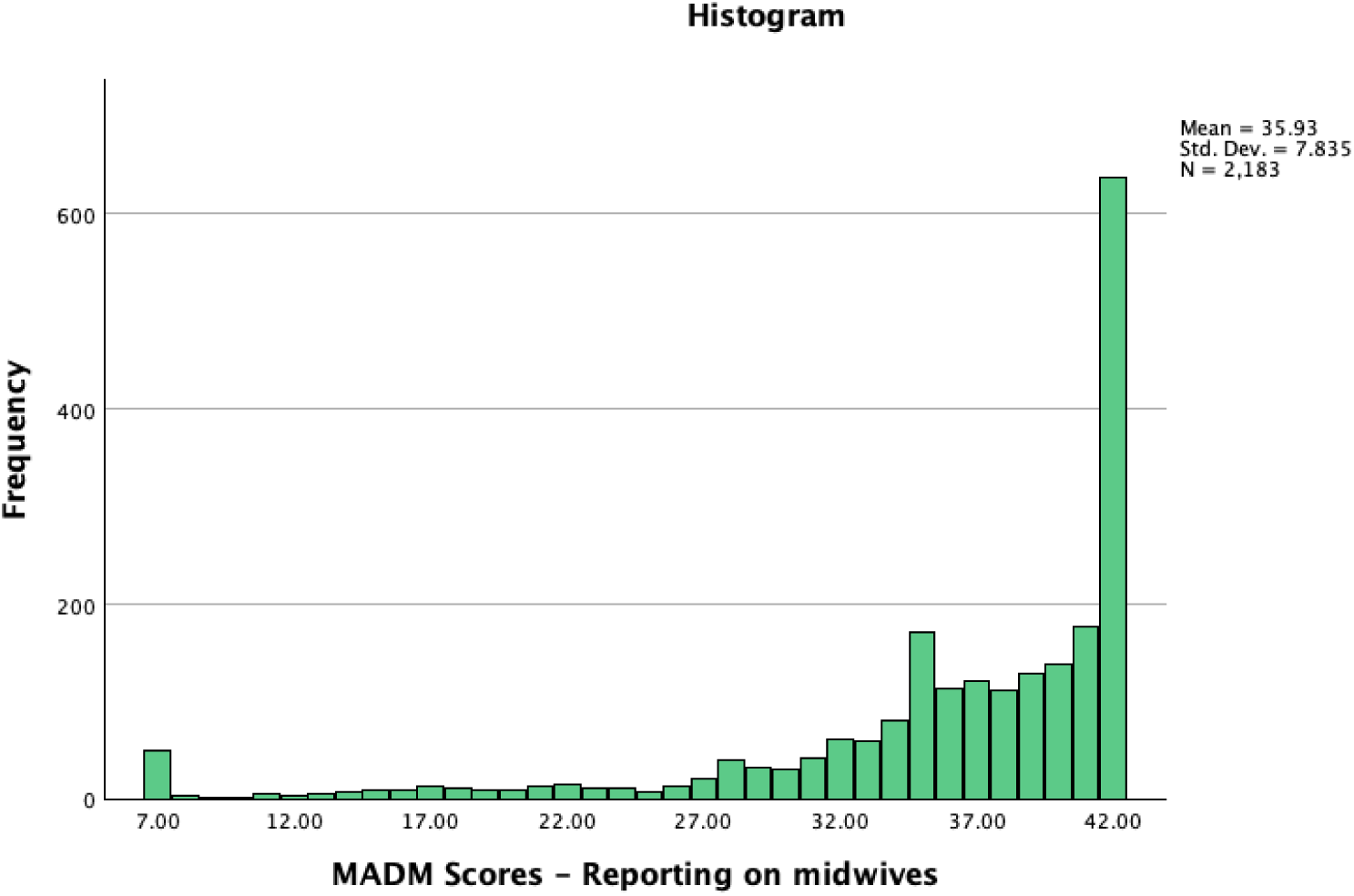
Distribution of MADM scores among participants reporting on midwives (n=1909 participants reporting 2138 autonomy experiences)

### Data analysis

The dataset was organized so that each row of data represented one autonomy experience. This means that for the 1660 respondents who completed the MADM scale twice to report on different care providers, the variables used in the analysis were copied over and we controlled for repeat observations referring to a single pregnancy in the multivariable models. Participants who experienced a pregnancy loss were excluded from analysis; their experiences are reported elsewhere (48).

We recoded MADM scores into two groups: those who scored at or above the 80^th^ percentile (defined as high autonomy) and those who scored below the 80^th^ percentile (defined as average or low autonomy). Given the different distributions of scores among participants who reported on interactions with midwives or physicians and, given that we wanted to compare autonomy experiences <u>within</u> rather than <u>across</u> the two models of care, we opted to create health care provider-specific cut off scores. The cutoff score for physicians was 35 and for midwives 42 (corresponding to scores at or above the 80th percentile).

We report frequencies and proportions to describe our sample. To assess the association between high autonomy and participant characteristics, circumstances, and contextual factors, we performed Pearson’s chi square analysis, stratified by health care provider type.

#### Multivariable modelling

To determine statistical significance and magnitude of difference, we calculated the odds of high autonomy and 95^th^ confidence intervals (95% CIs) among subgroups using binary logistic regression analysis. We controlled for year of pregnancy because temporal trends in this dataset indicated an increase in autonomy over the years of data collection among participants, especially those reporting on physicians (see Figure 3) and because some participants reported on pregnancies during the COVID-19 pandemic which might have affected their care. We controlled for the pregnancy that participants chose to report on because individuals with previous pregnancies are likely to have different expectations and experiences of decision-making in pregnancy compared to people who are going through this experience fopr the first time. We also controlled for repeat observations to account for response patterns among persons who submitted two MADM scores. A mixed effects model was considered (where participants who submitted two versus one autonomy experience were entered as a random effect), but no clustering of scores based on the number of autonomy experiences was detected in an intercept-only mixed effects model. In the logistic regression models, the unit of analysis was the autonomy experience whereas the person was the unit of analysis for the descriptive and bivariate analyses.

**Figure 3:**
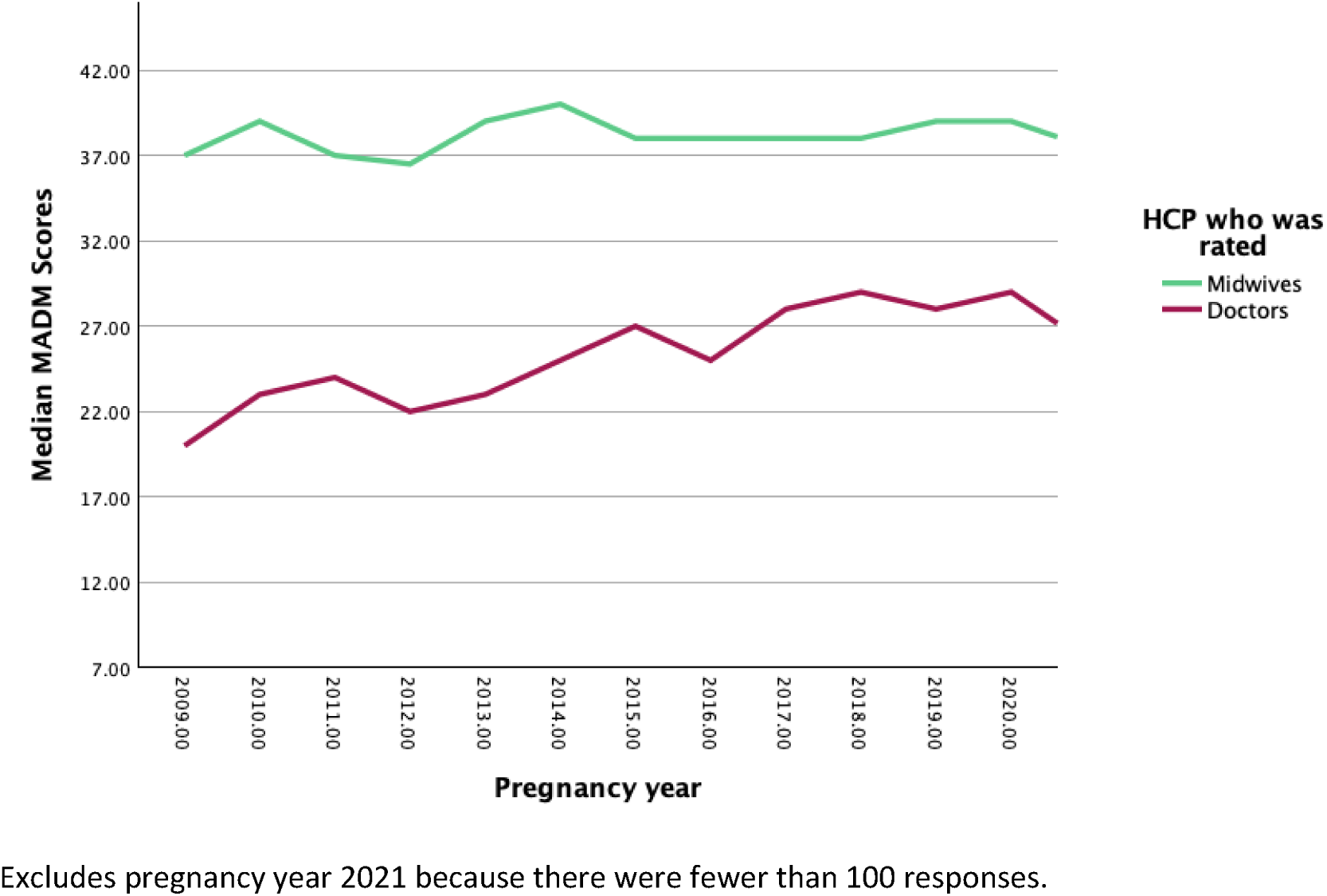
Temporal changes in autonomy score, by health care provider type (n=5295)

We ran separate logistic regression models for each independent variable and for people who rated physician-led versus midwife-led interactions. We also report odds and 95^th^ % confidence intervals for all health care providers combined. For these analyses, we defined high autonomy as scores at or above the 80^th^ percentile (which corresponded to a score of 39 or higher when looking at the distribution of MADM scores for all providers combined).

## RESULTS

### Internal consistency of MADM scale in the RESPCCT Study

Internal consistency reliability of the different language versions of the MADM scale was high, ranging from 0.89-0.97 (see Additional File 2). Most participants completed the English version of the scale (n=4828), and 478 completed the French version. We received 33 completed scales in Arabic, 12 in Spanish, 6 in Punjabi, 11 in simplified Chinese, 21 in traditional Chinese, and none in Inuktitut.

### Sample

One in three respondents (34.7 %) resided in Ontario during pregnancy, 22.5 % in British Columbia, 18.0 % in the Prairie provinces, 12.3 % in Quebec, 11.0 % in the Atlantic provinces and 1.5 % in the Territories. Participant characteristics are described in Table 2. Most respondents (87.8 %) said it was extremely important or very important to lead decisions about their pregnancy, birth, and/or baby.

**Table 2:** Association between maternal characteristics, and high autonomy.

|  | Unit of analysis is person (n=5389) |  |  | Unit of analysis is MADM score (n=7040) |  |  |
| --- | --- | --- | --- | --- | --- | --- |
|  | n (%) | n (%)<br>High<br>autonomy<br>Reporting on<br>physicians | n (%)<br>High<br>autonomy<br>Reporting on<br>midwives | Odds ratios<br>(95% CIs)<br>Reporting on<br>physicians<br>(n=4693) | Odds ratios<br>(95% CIs)<br>Reporting on<br>midwives<br>(n=2183) | Odds ratios<br>(95% CIs)<br>All providers (n=7049) |
| **Ethnoracial identity** |  | p=0.306 | p=0.772 |  |  |  |
| Indigenous | 288 (6.5) | 40 (20.1) | 16 (21.6) | NS | NS | **0.73 (0.59-0.90)** |
| Black | 139 (3.1) | 20 (20.6) | 12 (30.0) | NS | NS | NS |
| Middle Eastern | 100 (2.3) | 19 (23.5) | -- | NS | NS | **0.57 (0.39-0.83)** |
| South Asian | 91 (2.1) | 8 (15.7) | 10 (25.6) | NS | NS | NS |
| East Asian | 156 (3.5) | 32 (31.1) | 13 (26.0) | NS | NS | NS |
| Latine or Hispanic | 101 (2.3) | 18 (29.0) | 12 (30.8) | NS | NS | NS |
| Other | 80 (1.8) | 13 (26.5) | 10 (33.3) | NS | NS | NS |
| White | 3469 (78.4) | 511 (24.7) | 418 (30.8) | Reference | Reference | Reference |
| Gender identity |  | p=0.734 | p=0.877 |  |  |  |
| Gender minority | 115 (2.7) | 17 (25.8) | 13 (28.9) | NS | NS | NS |
| Cis Gender | 4152 (97.3) | 610 (23.9) | 462 (30.0) | Reference | Reference | Reference |
| Age at start of pregnancy |  | p < 0.001 | p=0.698 |  |  |  |
| < 25 years | 478 (10.8) | 53 (15.1) | 36 (33.0) | **0.58 (0.43-0.78)** | NS | **0.47 (0.36-0.62)** |
| 25-34 | 2996 (67.6) | 461 (25.3) | 346 (30.5) | NS | NS | NS |
| 35 and over | 959 (21.6) | 152 (27.3) | 114 (29.0) | Reference | Reference | Reference |
| Education |  | p < 0.001 | p=0.904 |  |  |  |
| Some high school or high school completed | 315 (7.5) | 41 (17.7) | 20 (28.6) | **0.67 (0.49-0.91)** | NS | **0.37 (0.27-0.50)** |
| Some college or apprenticeship completed | 999 (23.7) | 133 (20.3) | 94 (28.8) | **0.67 (0.54-0.83)** | NS | **0.72 (0.61-0.85)** |
| Undergraduate degree | 1425 (33.7) | 205 (24.1) | NS | NS | NS | NS |
| Grad degree/professional degree | 1485 (35.2) | 237 (28.1) | 186 (29.7) | Reference | Reference | Reference |
| **Disability during pregnancy** |  | p=0.134 | p=0.600 |  |  |  |
| Yes | 333 (7.8) | 43 (19.8) | 30 (27.8) | NS | NS | **0.77 (0.61-0.98)** |
| No | 3959 (92.2) | 585 (24.4) | 452 (30.2) | Reference | Reference | Reference |
| **Relationship status during pregnancy** |  | p=0.233 |  |  |  |  |
| Single | 121 (2.8) | 16 (18.6) | -- | Reference | -- | Reference |
| Married or partnered | 4211 (97.2) | 622 (24.2) | 485 (30.7) | NS | -- | **2.48 (1.51-4.07)** |
| **Immigration history** |  | p=0.013 | p=0.015 |  |  |  |
| Born in Canada | 2921 (66.1) | 394 (22.0) | 343 (31.6) | NS | NS | NS |
| Immigrated within 3 years | 195 (4.4) | 33 (25.6) | NS | NS | **0.34 (0.15-0.77)** | **0.60 (0.37-0.97)** |
| Immigrated within 4-6 years | 132 (3.0) | 23 (30.7) | 21 (38.9) | Reference | Reference | Reference |
| Immigrated > 6 years ago | 1170 (26.5) | 198 (27.5) | 120 (27.5) | NS | NS | NS |
| **Substance use during pregnancy** |  | p=0.313 | 0.479 |  |  |  |
| Yes | 181 (4.4) | 24 (20.5) | 20 (34.5) | NS | NS | **0.68 (0.49-0.95)** |
| No | 3979 (95.6) | 596 (24.6) | 451 (30.1) | Reference | Reference | Reference |
| **Ability to meet financial obligations** |  | **p < 0.001** | **p=0.028** |  |  |  |
| Income not enough | 340 (8.0) | 32 (13.9) | 28 (28.6) | **0.49 (0.36-0.68)** | NS | **0.47 (0.36-0.62)** |
| Income Enough | 1724 (40.3) | 232 (21.1) | 157 (26.4) | **0.73 (0.62-0.86)** | **0.78 (0.63-0.97)** | **0.64 (0.56-0.72)** |
| Income more than enough | 2210 (51.7) | 362 (28.4) | 298 (32.8) | Reference | Reference | Reference |
| **# of social services needed during pregnancy** |  | **p=0.192** | **p=0.007** |  |  |  |
| None | 2809 (80.4) | 429 (25.7) | 363 (32.8) | Reference | Reference | Reference |
| 1 | 501 (14.3) | 63 (21.9) | 48 (24.0) | NS | **0.63 (0.46-0.88)** | **0.80 (0.65-0.97)** |
| 2 or more | 185 (5.3) | 29 (20.7) | 6 (16.2) | NS | **0.35 (0.16-0.79)** | **0.33 (0.22-0.50)** |
| **Was told by HCP that pregnancy is high risk** |  | **p=0.163** | **p=0.003** |  |  |  |
| Yes | 1053 (19.5) | 222 (25.0) | 27 (18.4) | NS | **0.59 (0.41-0.86)** | **0.46 (0.39-0.54)** |
| No | 4336 (80.5) | 566 (22.7) | 525 (29.8) | Reference | Reference | Reference |
| **Pre-pregnancy Body Mass Index (BMI)** |  | p=0.643 | p=0.780 |  |  |  |
| < 18.5 | 155 (3.7) | 29 (27.9) | 15 (30.6) | NS | NS | NS |
| 18.5-29.9 | 2151 (51.2) | 445 (23.9) | 391 (30.5) | Reference | Reference | Reference |
| 30.0 and higher | 1039 (24.7) | 143 (23.9) | 68 (28.2) | NS | NS | **0.78 (0.66-0.91)** |
| **Day-to-day discrimination** |  | **p < 0.001** | **p=0.014** |  |  |  |
| None (score of 0) | 2009 (49.4) | 331(27.8) | 263 (33.4) | Reference | Reference | Reference |
| Low (score of 1 or 2) | 1118 (27.5) | 152 (22.6) | 112 (26.2) | 0.85 (0.71-1.03) | **0.70 (0.55-0.90)** | **0.84 (0.72-0.97)** |
| Moderate/high<br>(score of 3 or higher) | 938 (23.1) | 112 (18.6) | 86 (27.1) | **0.64 (0.52-0.79)** | **0.73 (0.56-0.95)** | **0.72 (0.61-0.84)** |

Results are based on binary logistic regression models, and controlled for year of pregnancy, repeat observations (i.e. persons who submitted two MADM scores), and the pregnancy reported on (first, second, third or fourth or higher). Statistics are only displayed for groups with 30 or more participants. NS= Not significant

5389 participants submitted 7049 MADM scores. The mean MADM score was 28.6 (standard deviation = 10.7) and the median 31 (5^th^, 95^th^ percentile: 8, 42). Large increases in scores over time were observed for those who completed MADM in reference to physicians (obstetricians and family physicians). Scores for midwifery clients mostly stayed the same over time (see Figure 3). Almost all (95.0%) who completed the MADM scale for the first provider reported on the provider type who was leading their pregnancy care.

### Association between maternal characteristics and high autonomy - all providers combined

When responses from all participants were included (ie. not stratified by provider type), many groups reported significantly lower odds of high autonomy. For example, we found significantly reduced odds of high autonomy among Indigenous (AOR=0.73; 95 % CI: 0.59-0.90) and Middle Eastern (AOR=0.57; 95 % CI: 0.39-0.83) compared to white participants and among individuals who reported one or more disabilities during pregnancy (AOR=0.73; 95 % CI: 0.59-0.89) (see Table 2). Participants who reported low compared to no discrimination had significantly lower odds of high autonomy (AOR=0.81; 95 % CI: 0.68-0.95). Loss of autonomy was more pronounced when comparing people with moderate and high levels of day-to-day discrimination with those who experienced no discrimination (AOR=0.69; 95 % CI: 0.57-0.83).

### Maternal characteristics and high autonomy -interactions with physicians

The following respondents who reported on physician interactions reported significantly lower odds of high autonomy: individuals who were under 25 years of age or younger during pregnancy (compared to those 35 or older), those without a university degree (compared to a graduate or professional degree), and participants who did not have enough income or just enough income to meet monthly financial obligations (compared to those who had more than enough income). Characteristics that were associated with the largest gaps in autonomy were: age < 25 years, moderate to high discrimination, low income and lower educational attainment.

### Maternal characteristics and high autonomy -interactions with midwives

Among respondents who rated midwives the following maternal characteristics were associated with significantly lower odds of high autonomy: Being a newcomer to Canada (compared to having been in the country between 4-6 years), having enough income (compared to more than enough income to meet monthly financial obligations), needing social services during pregnancy (compared to not needing services), being told by their health care provider that their pregnancy is high risk, and experiencing low or moderate/high (compared to no) discrimination. Maternal characteristics that were associated with the largest gaps in autonomy were: being a newcomer to Canada and needing two or more social services during pregnancy (see Table 2).

### Association between context of care and high autonomy

Several context of care variables were associated with increased odds of high autonomy, especially among respondents who rated physicians: Those who rated interactions with physicians and answered yes to the question *‘Did you get prenatal care as early as you liked?’* were more than twice as likely to report high autonomy than people who answered no to the statement. Those who rated physician-led care and *felt that they had enough time during prenatal visits* were more than six times as likely to report high autonomy compared to people who disagreed with the statement. Participants who rated interactions with physicians and were *easily able to find their preferred type of prenatal provider* were nearly 4 times as likely to report high autonomy compared to people who could not easily find their preferred provider. The odds of high autonomy significantly increased when participants reported more optimal access conditions (see Table 3) but these associations were only significant for participants who rated physicians.

**Table 3:**
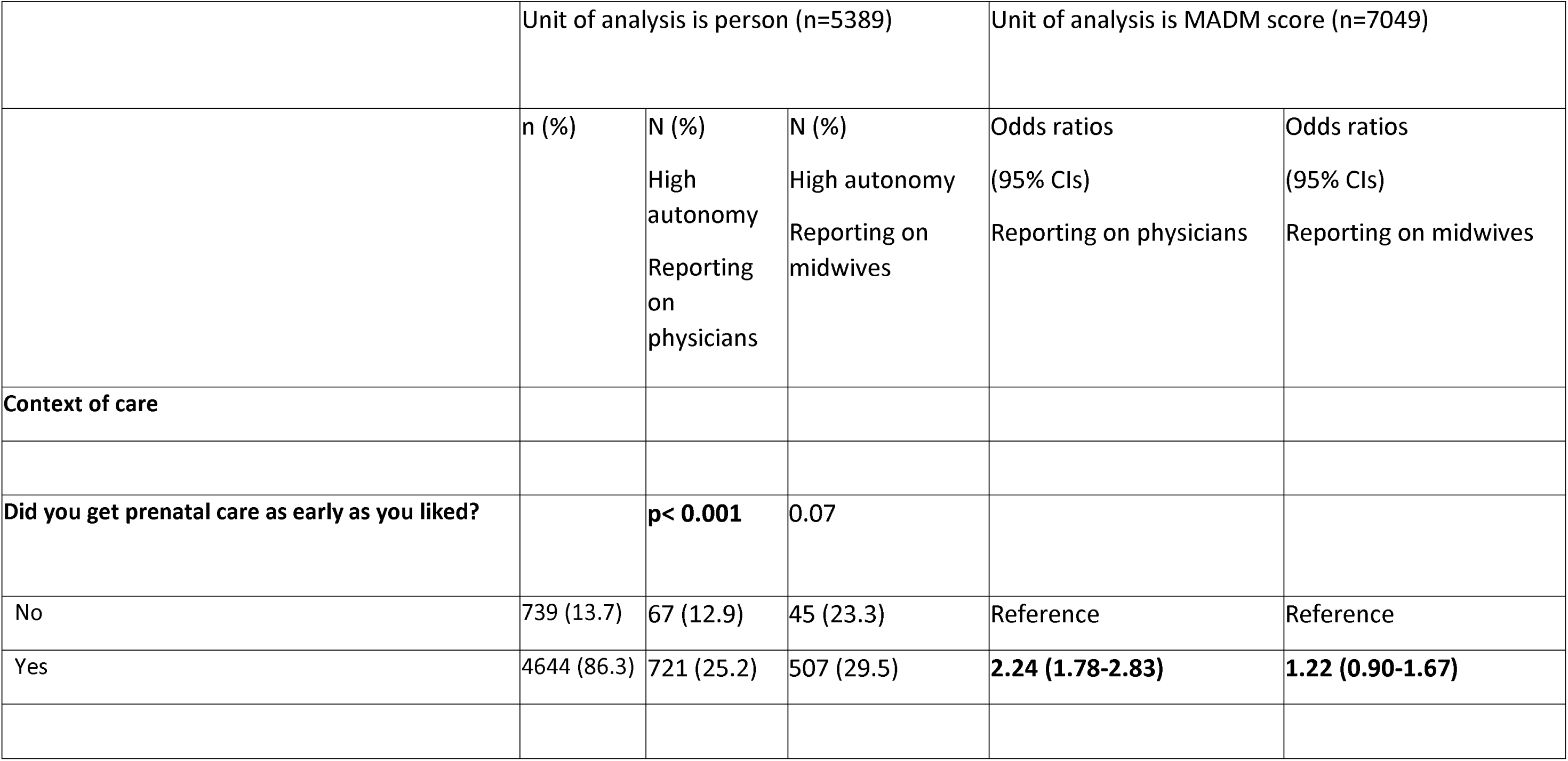
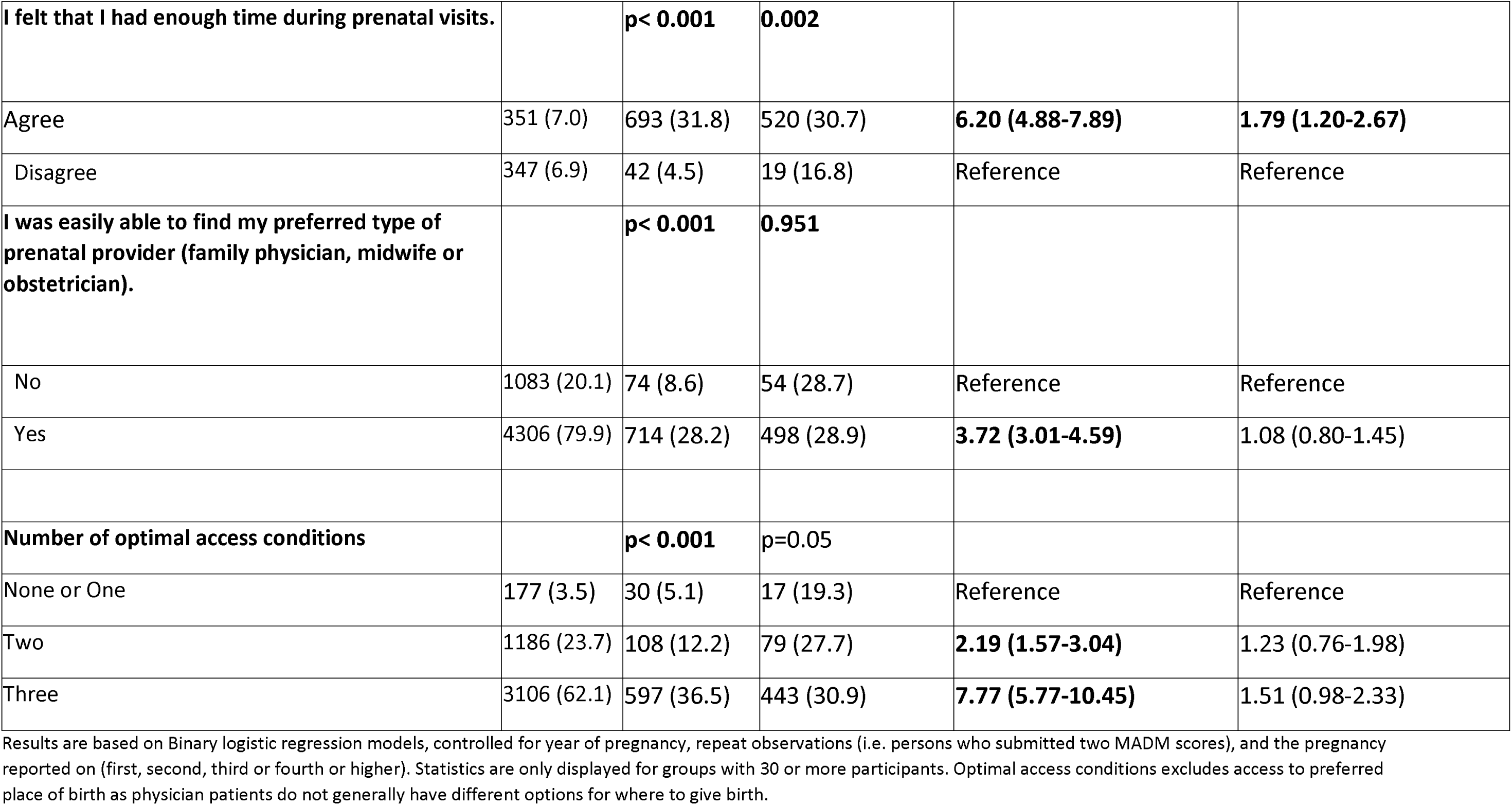
Association between contextual factors and high autonomy.

## DISCUSSION

This is the first national study in Canada to examine experiences of agency in decision-making during pregnancy across a broad cross-section of the childbearing population. Strengths of the study include the large and diverse sample of underrepresented women; the application of validated measures to assess autonomy and day-to-day discrimination; the community-driven approach to item generation and selection, data collection, analysis and interpretation; and our ability to compare our findings with other studies that used the same measure of autonomy. While most respondents preferred to lead decisions about their care, there were significant disparities in their ability to exercise autonomy by personal characteristics and level of day-to-day discrimination. As hypothesized, minoritized, and marginalized individuals report greater loss of autonomy.

When analyzing the full sample, ie. not stratified by provider type, Indigenous and Middle Eastern individuals had significantly lower odds of autonomy, compared to white respondents. These findings align with ubiquitous reports of anti-Indigenous racism in Canadian healthcare and lack of agency among racialized childbearing people (49–52), including Indigenous women being harmed disproportionately by relocation for birth (also referred to as maternal evacuation policy), and referrals to child protective services (birth alerts). The characteristics most strongly associated with loss of autonomy were indicators of socio-economic standing (such as the inability to meet financial obligations, less education, and need for social services)(53). The association between lower levels of education, financial stress and lower MADM scores has been documented by other Canadian authors (23,35,54). Aligned with our results, others have found significantly lower autonomy among migrants(45).

In the current study, the odds of high autonomy were significantly lower among participants with pregnancy complications who were reporting on interactions with midwives. This aligns with findings from a survey study from Iceland where the majority of perinatal care is delivered by midwives (38), a survey study from Canada, and a qualitative study from Switzerland that showed perinatal providers were less likely to support autonomy when complications arise (23,39). The finding that loss of autonomy was more common among those with higher BMI (all providers combined) adds to a growing body of evidence on weight stigma and discrimination in Canadian healthcare (55).

Higher levels of day-to-day discrimination were significantly associated with loss of autonomy across all three models of care (all providers combined and stratified by provider type). It is important to include validated measures of structural factors like discrimination in perinatal studies to avoid using race and other personal identifiers as proxies for racism (42,43). If we assume that it is solely a childbearing person’s identity that links to lower autonomy, as opposed to recognizing structural and historical drivers of inequities in health care interactions, we can perpetuate stereotypes and undermine the strength-based approach that is emerging in public health research (42,43). Other published studies that used the MADM scale did not measure or did not find significant associations with autonomy and younger age, having one or more disabilities during pregnancy, relationship status, reporting substance use during pregnancy and experiences of day-to-day discrimination. Hence, the current analysis provides new evidence about the link between autonomy and these participant characteristics.

In the current study, access to their preferred provider was linked to high autonomy. Likewise, participants who received care from a chosen physician or midwife in Hungary reported a median score of 27 and 31 compared to a score of 21 among those who did not have access to a chosen provider (56). Our study highlights that enough time during prenatal visits was strongly associated with high autonomy. This finding is supported by research from British Columbia (BC), using the same scale to measure autonomy. Childbearing persons from BC who held back questions during prenatal appointments because their provider seemed rushed reported significantly lower MADM scores (36).

In addition, participants with prenatal appointments less than 15 minutes reported median MADM scores of 23 compared to 36 for those with appointments lasting between 16-30 minutes. The highest MADM scores were reported for people with appointment lasting more than 30 minutes (35).

Research from the UK also supports the link between autonomy and length of appointments (57).

### Model of care

Autonomy scores among those reporting on interactions with midwives were more than 10 points higher than other HCP types (see Figure 3), a finding that is supported by several studies that examined MADM scores across healthcare provider groups (17,23,35,44,45,56,58). In the current study, Indigenous and Middle Eastern respondents were the least likely to report prenatal midwifery care: 22.9 % of Indigenous people and 15.2 % of Middle Eastern participants reported prenatal midwifery care, compared to 38.5 % of white participants. Other groups who were significantly less likely to have midwifery care were: those 25 and younger, people with lower educational attainment, people with one or more disabilities during pregnancy, single parents and people in need of social services (see Table 4). Marginalized and minoritized women in the current study were less likely to report having prenatal midwifery care, a finding that fills a gap in the literature as previous studies were mostly focused on exploring barriers to midwifery care for uninsured women or those with low socio-economic standing (59,60). Fortunately, our findings that autonomy scores for physicians are improving over time (see Figure 1) suggests that public and health professional education around person-centred care may be improving the overall experience of decision-making, regardless of type of provider during pregnancy in Canada.

**Table 4:** Differences in prenatal midwifery care, by maternal characteristics (n= 5389)

|  | % who had a midwife as their primary prenatal provider * | p |
| --- | --- | --- |
| Ethnoracial identity |  | < 0.001 |
| Indigenous | 69 (22.9) |  |
| Black | 40 (28.4) |  |
| Middle Eastern | 16 (15.2) |  |
| South Asian | 40 (42.1) |  |
| East Asian | 48 (29.6) |  |
| Latine or Hispanic | 37 (35.9) |  |
| Other | 32 (39.0) |  |
| White | 1366 (38.5) |  |
| Gender identity |  | 0.758 |
| Cis Gender | 1528 (36.8) |  |
| Gender minority/gender diverse | 44 (38.3) |  |
| Age at start of pregnancy |  | < 0.001 |
| < 25 years | 105 (22.0) |  |
| 25-34 | 1130 (37.7) |  |
| 35 and over | 386 (40.3) |  |
| Education |  | < 0.001 |
| Some high school or high school completed | 66 (21.0) |  |
| Some college or apprenticeship completed | 331 (33.1) |  |
| Undergraduate degree | 552 (38.7) |  |
| Graduate degree/professional degree | 618 (41.6) |  |
| Disability |  | 0.030 |
| No | 1488 (37.6) |  |
| Yes | 105 (31.5) |  |
| Relationship status |  | < 0.001 |
| Single | 23 (19.0) |  |
| Married or partnered | 1570 (37.3) |  |
| Immigration history |  | 0.168 |
| Born in Canada | 1072 (36.7) |  |
| Immigrated within 3 years | 58 (29.7) |  |
| Immigrated within 4-6 years | 54 (40.9) |  |
| Immigrated > 6 years ago | 436 (37.3) |  |
| Substance use during pregnancy |  | 0.130 |
| No | 1488 (37.4) |  |
| Yes | 57 (31.5) |  |
| Ability to meet financial obligations |  | < 0.001 |
| Income more than enough | 909 (41.1) |  |
| Income Enough | 586 (34.0) |  |
| Income not enough | 95 (27.9) |  |
| # of social services needed during pregnancy |  | < 0.001 |
| None | 1106 (39.4) |  |
| 1 | 189 (37.7) |  |
| 2 or more | 39 (21.1) |  |

\*The type of care provider most directly involved in my PRENATAL care (i.e. visits for check-ups and advice on your pregnancy)

#### IMPLICATIONS FOR PRACTICE AND POLICY

In this national study, using patient-reported experience measures, we show that disparities in autonomy exist in Canada and are more pronounced for people with less privileged social positions. In a healthcare system where people are purported to be treated the same irrespective of their social identities and circumstances, we would expect to find no differences in autonomy experiences. The fact that these differences were observed irrespective of which type of care provider people reported on suggests that the impact of structural inequities in the overall health system transcends differences in models of prenatal care in Canada.

The Canadian Medical Association Code of Ethics (61) asks physicians to ‘empower the patient to make informed decisions regarding their health by communicating with and helping the patient navigate reasonable therapeutic options to determine the best course of action consistent with their goals of care’ (p. 5) and to ‘respect the decisions of the competent patient to accept or reject any recommended assessment, treatment, or plan of care (p.5). Registered midwives in Canada are expected to provide each client with information regarding options for relevant treatments, procedures, tests and medications throughout their care (62). The British Columbia College of Nurses & Midwives (BCCNM) Informed Choice Policy delineates the types of information that all midwives are expected to provide: “what is being proposed/offered and its risks/benefits; any alternatives to what is being proposed/offered and their risks/benefits; relevant research evidence including any deficiency of clear evidence; what would happen if no treatment/procedure/test/medication is chosen; and relevant community standards of care and practices. The midwife must also make reasonable efforts to ensure that the client has adequate opportunity and time to engage in the informed choice process.” (63)(p.1). However, despite clear standards that support a pregnant person’s agency to make decisions about their care, a chasm between guidelines and reality exists, and myriad system- and individual level factors influence whether, how much, and *for whom* autonomy is enabled.

Our analysis elicited several instructive findings on facilitators of autonomy, including modifiable health systems factors, such as adequacy of time during prenatal appointments, access to preferred providers, and timeframe of onset of prenatal care. Policies that respond to these findings could mandate adequate reimbursement, across models of care, for longer prenatal appointments, and/or dedicated, culture and trauma informed perinatal care staff who can help families to understand and navigate access to community-specific options for care.

### Barriers & Facilitators to enabling autonomy in decision-making during pregnancy

The options that pregnant people are presented with and the degree to which health care providers support autonomy in decision-making is influenced by many factors, including the attitudes of health care providers (HCPs) (64–66). HCPs in Switzerland questioned the decision-making capacity of birthing people and described factors that advance and limit autonomy (67). Facilitators included: building trust during antenatal appointments, using tools to improve communication, giving sufficient time to make decisions, and postnatal debriefing. Limiting factors included reduced knowledge about reproductive rights, and lack of birth preparation or decisional capacity (among childbearing people). Other barriers to autonomy that emerged during interviews were conflicts among practitioners due to different perspectives about autonomy, lack of continuity of care weakening trust between providers and patients, and structural barriers such as resource limitations, hospital guidelines/protocols, medical hierarchy, and health politics that may override personal choices of birthing people (67).

Even well-intentioned attempts to offer choices can negatively impact reproductive autonomy and informed decision-making. For example, in Sangster and Lawson’s qualitative study (68), participants described experiences that threatened their ability to make informed decision about prenatal testing for Down Syndrome: routinization of screening (i.e. being screened without offering it as a choice and HCP not explaining that screening is optional); HCPs equating an openness to screening with an openness to testing and termination; and misunderstanding and miscommunicating about the probability of different outcomes.

Most importantly, our study supports findings from recent studies on experiences of health care among various marginalized populations. Childbearing people with disabilities in Ontario reported ableist assumptions from healthcare providers, and limited ability to make decisions about their care (28). People with disabilities feared judgement and discrimination from healthcare providers and reported that providers lacked awareness of how to accommodate them (69). Women with high pre-pregnancy body mass index (BMI) were more likely to report disrespectful care and loss of autonomy compared to people of normal BMI in a pan-Canadian study (54). Empirical evidence about factors that improve perinatal experiences is emerging in Canada, including midwifery care, longer prenatal appointments, and assuring cultural safety, such as honoring Indigenous rights to incorporate culture and ceremony into birth care (70,71). While addressing structural inequities requires multisectoral, long-term engagement and commitment, our study confirms an urgent need to transform the education and preparation of health care providers to understand how to engage in inclusive, equitable, and person-centred decision-making processes.

See Box 1 for recommendations to improve autonomy in perinatal health care decision-making.

### Limitations and Future Research

In Canada among three distinct Indigenous populations, First Nations, Inuit, and Métis, there are over 600 communities. Each have unique cultures, languages, and histories shaped by their specific relationships to the land and the impacts of colonization, making it essential to recognize these differences to address their unique needs, policies and rights effectively. However, the small sample size would not allow for a nation-specific analysis. Similarly, the small subsample of women with disabilities prevented analysis of autonomy experiences based on the number and types of disabilities and using the full range of response options for gender, identity, age, and BMI.

Options for care that are available to pregnant people depend on many system-level and individual factors, including geography, income, access to transportation, health status, health literacy, and the experience and comfort of HCPs. In the current study we measured some but not all these factors. Additional unmeasured factors that might influence perceptions of autonomy include individuals’ capacity to engage in decision making; pre-existing cultural expectations about the rights to choice; and the gender of prenatal providers, which has been shown to affect communication (72). Future studies ought to examine the relationship between autonomy and specific aspects of care such as continuity of care, which was linked to higher MADM scores in a study from Australia (7,13).

## Conclusion

We found that loss of autonomy was more common among respondents with marginalized sociodemographic identities and circumstances and more pronounced for participants who reported higher levels of discrimination. We identified several modifiable factors that appear to promote autonomy in pregnancy, including midwifery care, early entry into prenatal care, and the ability of pregnant people to access their preferred type of provider. These modifiable factors were especially important for those who reported on interactions with physicians. Our findings draw attention to the importance of having enough time to support informed decision-making across models of care and subpopulations; expansion of access to midwifery care for marginalized individuals; as well as universal attention to conversations between HCPs and pregnant people about their preferences and expectations for leading decisions. These structural factors should be seen as reproductive rights, alongside the right to make autonomous decisions during pregnancy, birth and the postpartum period (73,74). To date, the discourse about reproductive autonomy has been narrowly focused on controlling, assisting with and avoiding reproduction (e.g. forced sterilization, fertility services and abortion) and needs to be expanded to include all perinatal decisions and experiences (75,76). Doing so will likely improve health outcomes and experiences across communities regardless of marginalized or minoritized identity.

Box 1. Recommendations to improve autonomy in perinatal decision-making

*EDUCATION*

1. Provide information to pregnant individuals about their rights and the recourse available to them if their rights are not respected.
2. Enhance curricula to support acquisition of person-centred decision-making competencies across perinatal health professions.
3. Mandate ongoing, continuing interprofessional education on how to support person-centred decision-making during pregnancy and childbirth.
4. Systemize post-secondary training programs as well as continuing professional development opportunities on the topics of historical and structural racism, disparities, disability, trauma-informed care, Indigenous cultural safety and respectful care.

PRACTISE

1. Use a systematic process to center the pregnant person’s preferences for care throughout the course of care. https://www.birthplacelab.org/decision-making-tool/
2. Communicate clearly to patients that they can engage in decisions, and encourage them to do so. Whenever possible encourage pregnant women, families and key support persons to ask questions, discuss and clarify key expectations, and their preferences for care
3. Check their personal and cultural preferences for roles in decision-making. Communicate with transparency about any potential barriers to achieving their care preferences.
4. Provide pregnant people with adequate time to consider available options, and build in the time they need to decide on a course of action.
5. Introduce options for care using various communication methods (ie verbal, written, visual) and revisit decisions made.
6. Avoid signaling power imbalances, including attending to physical positioning during conversations and body language.
7. Support informed choice discussions by using lay language that can be understood by all involved. Increase access to language interpreters for medical care including access to Sign Language interpreters and offer information in lay language/multiple languages that can be helpful for many persons (low literacy, language barriers, feeling overwhelmed or exhausted, sensory or auditory processing challenges, etc.).
8. Enquire whether people who enter care found their preferred type of provider or options for perinatal care. If the answer is no, be aware that expectations of autonomy might be different as a result and more effort might be required to close the gap.
9. Engage in continuous professional development to enhance competencies in cultural safety, supporting person-led decisions, and interprofessional communication and collaboration.
10. Acknowledge, identify, and elevate attention to health systems factors that contribute to inequities in peoples experience.

HEALTH SYSTEM/HOSPITALS

1. Create clinic processes, staffing models, physical spaces and program budgets that adequately support the dedicated time needed to provide respectful, relationship-based care.
2. Support expansion of culture-centered care via Indigenous midwives, (e.g. reserved privileging spots for Indigenous midwives, salaried positions etc.).
3. For Indigenous families, establish facility-level policies that honor the right to ceremony and rituals surrounding pregnancy and birth. Birth ceremonies are directly connected to the land, so recognizing the importance of birthplace (geographical location) and honoring the sacredness of birth is a significant way to recognize and implement Indigenous rights (77) and promote autonomy.
4. As part of continuous quality improvement, routinely measure the experience of decision-making during pregnancy and birth as an important indicator of quality perinatal care (see examples here: https://www.birthplacelab.org/measures-and-metrics/).
5. In collaboration with women and communities, co-develop strategies to improve autonomous decision-making among those at increased risk for adverse perinatal experiences.
6. Establish mechanisms within facilities or health authorities for childbearing people to submit reports of rights violations during birth and be transparent about mechanisms for redress.
7. Identify independent bodies to lead full and fair investigations into allegations of loss of autonomy and mistreatment during pregnancy and childbirth. Ensure that victims of rights violations are provided accountability and remedies by both government, regulatory bodies, facility, and/or non-state actors.
8. Continue to seek out opportunities to expand knowledge of health care rights and to advocate for the expansion of access to respectful and inclusive care options
9. Publish facility and provincial standards and commitments to supporting the rights to choose their healthcare providers and approach to health services, including the right to be informed about and have access to their preferred and culturally-responsive options for care, in the settings of their choice.
10. Work towards a national strategy to reduce structural inequities in provision of perinatal care.

## Data Availability

We will make the dataset available in the digital repository Dataverse, without restrictions (https://borealisdata.ca/dataverse/ubc), if and when the paper is accepted for publication.

## Additional Files

1. Additional file 1
  a. Title: Description of variables included in the analysis
  b. Format: Word Document
  c. Description: Description of variables that were included in the analysis.

2. Additional File 2
  a. Title: Internal Consistency Reliability of different language versions of MADM Scale
  b. Format: Word Document
  c. Description: The number of participants who completed the MADM scale in the various language versions and the internal consistency reliability of the different language versions of the MADM scale.

